# Timing of Medication Treatment in Children 3-5-Years-old with ADHD: A PEDSnet Study

**DOI:** 10.1101/2025.05.28.25328526

**Authors:** Yair Bannett, Ingrid Luo, Rodrigo Azuero-dajud, Heidi M. Feldman, Farah W. Brink, Tanya E. Froehlich, Holly K. Harris, Kristin Kan, Kate E. Wallis, Kaitlin Whelan, Lisa Spector, Christopher B. Forrest

## Abstract

**Importance:** Early identification and treatment of Attention-Deficit/Hyperactivity Disorder (ADHD) symptoms in preschool-age children is important for mitigating social-emotional and academic problems. Clinical practice guidelines recommend first-line behavior intervention before considering medication treatment for children 4-5-years-old.

**Objective:** To assess variation in rates of ADHD identification and rates and timing of medication treatment in children 3-5-years-old in primary care settings across eight US pediatric health systems and to identify patient factors associated with the time from diagnosis to prescription.

**Design:** Retrospective cohort study of electronic health records.

**Setting:** Primary care clinics affiliated with eight academic institutions participating in the PEDSnet Clinical Research Network.

**Participants:** Children 3-5-years-old seen in primary care between 2016-2023.

**Exposure:** ADHD diagnosis at age 4-5 years.

**Main Outcomes and Measures:** Outcomes: (1) rate of ADHD diagnosis; (2) rate of stimulant and non-stimulant prescription after diagnosis before age 7, (3) time from first ADHD-related diagnosis (including symptom-level diagnoses) to medication prescription. Independent variables: institution, year of diagnosis, patient age, sex, race/ethnicity, medical insurance, and presence of comorbidities.

**Results:** Of 712,478 children seen in primary care at ages 3-5 years, 9,708 (1.4%) received an ADHD diagnosis at age 4-5 years (range 0.5-3.1% across institutions). Of those with ADHD, 76.4% (n=7414) were male, 39.0% (n=3782) were White. Of 9,708 preschool-age children with ADHD, 68.2% (6624) were prescribed ADHD medications before age 7, 42.2% (n=4092) were prescribed medications within 30 days of the first documentation of an ADHD-related diagnosis (range 26.0-49.0% across institution). Asian (aHR 0.50, CI 0.38-0.65), Hispanic (aHR 0.75, CI 0.70-0.81), and Black (aHR 0.90, CI 0.85-0.96) children with ADHD were less likely to be prescribed medication early compared to White children. Older (aHR 1.64, CI 1.57-1.72), male (aHR 1.74, CI 1.11-1.24) and publicly insured (aHR 1.10, CI 1.04-1.17) patients were more likely to be prescribed medication early compared to younger, female and privately insured patients, respectively.

**Conclusion and Relevance:** Many preschool-age children with ADHD seen in primary care in 8 large pediatric health systems were prescribed medications at or shortly after the first documented diagnosis. Future analysis of clinical documentation is needed to understand the reasoning behind early prescription patterns.

## INTRODUCTION

Attention-Deficit/Hyperactivity Disorder (ADHD) is a highly prevalent neurodevelopmental disorder, estimated to affect 10% of US children.^1^ ADHD is increasingly being diagnosed in children before they enter school. A national survey in 2022 found that 2.4% of children ages 3-5 years had a diagnosis of ADHD.^2^ Preschool-age children with symptoms of ADHD are at risk for social-emotional problems and academic failure.^3,4^ Recognizing the early onset of ADHD, the American Academy of Pediatrics (AAP) subcommittee published updated evidence-based clinical practice guidelines for primary care management of ADHD in 2011 and re-affirmed in 2019. These guidelines include a separate set of recommendations for management of ADHD and ADHD symptoms in preschool-age children (ages 4-5 years).^5,6^ The guidelines recommend that primary care pediatricians (PCPs) start with parent training in behavior management and then consider medication treatment with methylphenidate as a second-line treatment, given stronger evidence for behavioral intervention than for medications in this age group.^7^

Most US children with ADHD are first diagnosed and treated by their PCP.^8–10^ However, to date, very few studies assessed primary care management of ADHD in this young age group.^11^ Survey-based studies found variation across patient subgroups in parent-reported ADHD diagnosis and treatment rates in preschool-age children.^2,12,13^ Prior claims- and electronic health record (EHR)-based studies assessed trends in prescription rates of stimulants in preschool-age children but did not assess the timing of treatment in relation to the initial diagnosis.^14–17^ Furthermore, information on variation in ADHD diagnosis and treatment across US health systems is limited, and evidence on the role of sociodemographic factors, including race/ethnicity, in contributing to disparities in diagnosis and treatment is mixed.^10,18–22^ To our knowledge, there are no large multi-site studies that leveraged clinical data from EHRs to assess rates of PCP diagnosis and treatment of ADHD in preschool-age children, including adherence to practice guidelines in the timing of medication treatment, and differences in clinical care across sociodemographic groups.

In this study, we assessed rates of ADHD identification and medication prescription, and timing of medication treatment in children 3-5 years-old in primary care settings across eight large US pediatric health systems. We evaluated patient factors associated with the time from first diagnosis to prescription.

## METHODS

We present our study in accordance with the Reporting of Observational Studies in Epidemiology (STROBE) reporting guidelines.^23^ This study was approved by a multi-site single Institutional Review Board led by Children’s Hospital of Philadelphia (protocol 60809).

### Setting and Data Source

Eight institutions that participate in PEDSnet, a national pediatric learning health system, provided EHR data for this study. Participating institutions included: Children’s Hospital of Colorado, Children’s Hospital of Philadelphia, Cincinnati Children’s Hospital, Lurie Children’s Hospital of Chicago, Nationwide Children’s Hospital, Nemours Children’s Health, Packard Children’s Hospital Stanford, Texas Children’s Hospital. Institutions extract data on a quarterly basis from source EHR systems and transform the data to the Observational Medical Outcomes Partnership (OMOP) data model (currently v5.7). Data are uploaded to the PEDSnet Coordinating Center at the Children’s Hospital of Philadelphia where they undergo rigorous data quality assessment. For this study, we extracted data from version v56 of the PEDSnet database on April 18, 2025.

### Study design and cohort selection

This was a retrospective cohort study of EHR data from all encounters (office/telehealth/telephone/administrative) of children ages 3-5 years seen between January 1, 2016, to December 31, 2023, in primary care clinics affiliated with eight academic institutions. We included children who were seen ≥2 times with at least 6 months of follow-up.

Primary care clinics were defined as having (1) ≥5% well-care visits among all visits, and (2) ≥1 childhood immunization procedure code (**eTable 1**). Subsequently, manual filtering removed care sites whose names were null or included the word ‘laboratory’.

We defined an ADHD-related encounter as an encounter with ADHD diagnoses or medication prescriptions (stimulants: methylphenidate, amphetamines; non-stimulants: alpha-agonists, atomoxetine). In a prior study, we found that PCPs frequently used symptom-level diagnoses of ADHD (e.g., hyperactivity) in children <6 years.^11^ Accordingly, we defined two types of ADHD diagnoses – (1) disorder-level (e.g., ADHD combined subtype), and (2) symptom-level (e.g., hyperactivity, inattention). Patients were included in the ADHD cohort if they received at least one disorder-level ADHD diagnosis between ages 4-5 years because guidelines recommend considering an ADHD diagnosis starting from age 4. ADHD medications prescribed after the initial diagnosis and before age 7 were included.

To capture PCP documentation of common developmental/behavioral comorbid conditions, as recommended by practice guidelines, we extracted for each patient any visit diagnosis of comorbid conditions (autism, anxiety, depression, disruptive behavior disorder, global developmental delay/intellectual disability, language delay/disorder, learning problem/disability, sleep problems) documented at or after the initial ADHD diagnosis.

**eTables 2-3** detail codesets for ADHD diagnoses, ADHD medications, and comorbid diagnoses.

### Study Outcomes and Measures

Primary Outcomes: (1) rate of ADHD diagnosis (disorder-level), (2) rate of stimulant and non-stimulant prescription after the initial diagnosis (disorder-level or symptom-level) and before age 7 years, and (3) time from initial ADHD diagnosis (disorder-level or symptom-level) to prescription (days).

Secondary Outcomes: (1) follow-up of prescribed patients within 60 days (ADHD-related encounter), and (2) documentation of comorbid conditions.

Patient sociodemographic and clinical characteristics, documented in the EHR, included age (continuous, at encounter of interest), sex, race/ethnicity (Asian/Non-Hispanic, Black/Non-Hispanic, Hispanic (irrespective of race), White/Non-Hispanic, Multiple Races/Non-Hispanic, Other/Non-Hispanic, Unknown), medical insurance at first ADHD-related encounter (private/public/other), presence of any developmental/behavioral comorbid condition, and number of primary care visits during the study period.

### Statistical analysis

Patient characteristics were summarized using frequencies (percentages) for categorical variables and median (interquartile range) for continuous variables. Values < 11 were reported as “<11” for confidentiality.

Time to first prescription was measured from the patient’s first ADHD diagnosis (disorder-level or symptom-level) after age 3 to before age 7. For descriptive analysis, time to prescription was categorized as: at or shortly after diagnosis (0-30 days), 1-6 months from diagnosis, and >6 months after diagnosis – a minimum timeframe recommended in the 2011 practice guidelines to pursue first-line behavioral treatment. We used inverse probability treatment weighting (IPTW) with cumulative incidence functions to visualize racial/ethnic differences while accounting for potential confounding by primary care utilization and care institution. Assumptions of positivity and between-subject independence were evaluated and met. We then fit multivariable Cox proportional hazards models to estimate associations between clinical and demographic predictors (i.e. EHR-recorded race/ethnicity, age at diagnosis, sex and insurance type) and time to prescription, adjusting for year of diagnosis, initial diagnosis type (disorder-level vs. symptom-level), comorbid conditions, primary care utilization (categorized as low [<5 visits], medium [5–10 visits], and high [>10 visits]), and institution. The proportional hazards assumption was tested and met.

Time from prescription to follow-up was visualized using cumulative incidence curves, presented overall and for each type of follow-up (in-person/telehealth) separately. Institutional variation in comorbidity documentation was shown using boxplots for any comorbidity documentation and for individual conditions.

In sensitivity analyses, we: (1) changed the definition of first ADHD-related encounter to include non-specific diagnosis codes of behavior problems (**eTable 4**), to assess the extent to which time to prescription is prolonged under the assumption that PCPs may use non-specific diagnoses (e.g., behavior concern) as the first indication of ADHD-related concerns. We used paired t-tests to compare time to prescription within the same patients under both definitions; (2) stratified time to prescription by initial ADHD diagnosis type (disorder-level vs. symptom-level) using cumulative incidence curves with log-rank tests, to test the hypothesis that when PCPs document symptom-level diagnoses first, time to prescription is longer than when they document disorder-level diagnoses first; and (3) repeat all analyses excluding institution E due to high rates of missing insurance data. All analyses were conducted using R version 4.3.2.

## RESULTS

Of 712,478 children seen in primary care at ages 3-5 years, 9,708 (1.4%) had an ADHD diagnosis (disorder-level) at ages 4-5 years (**Figure 1** shows study flowchart). Rate of ADHD diagnosis was variable across institutions (0.5-3.1%, **eTable 5**). Median age at first ADHD-related diagnosis was 5.3 years. Of 9,708 preschool-age children with ADHD, 7,414 (76.4%) were male and 3,782 (39.0%) were Non-Hispanic White; 6,624 (68.2%) were prescribed medications for ADHD, including stimulants (77.5% of prescribed children), non-stimulants (16.7%), and both (5.8%). Rate of ADHD medication prescriptions varied across institutions (44.1-74.1% of children) (**Table 1**).

**Figure 1.**
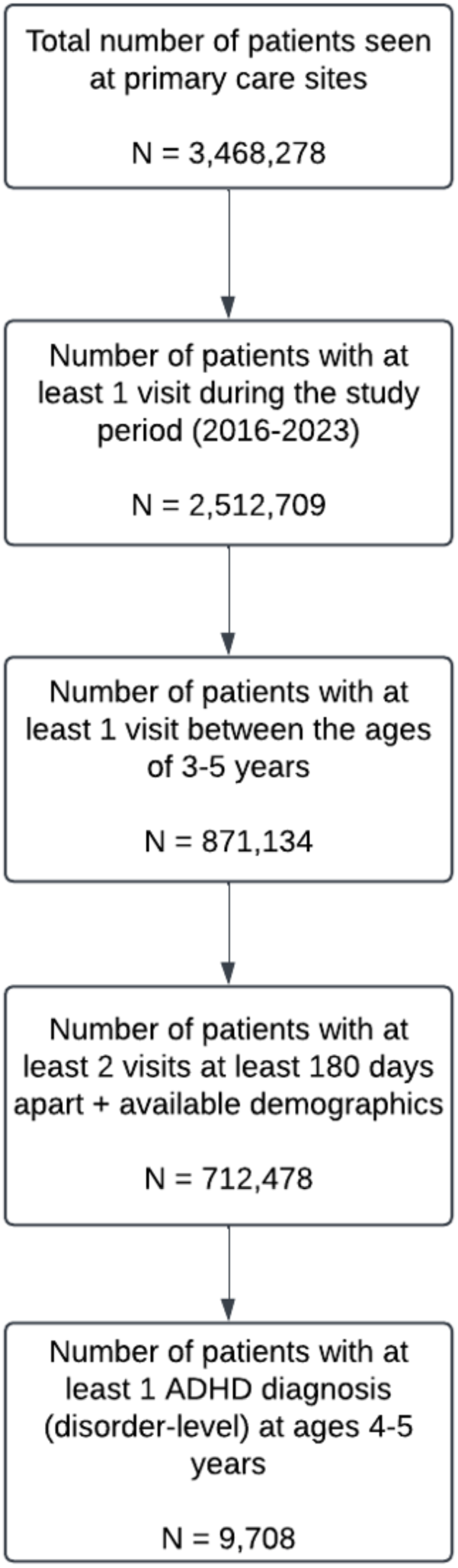
Study flowchart.

**Table 1.**
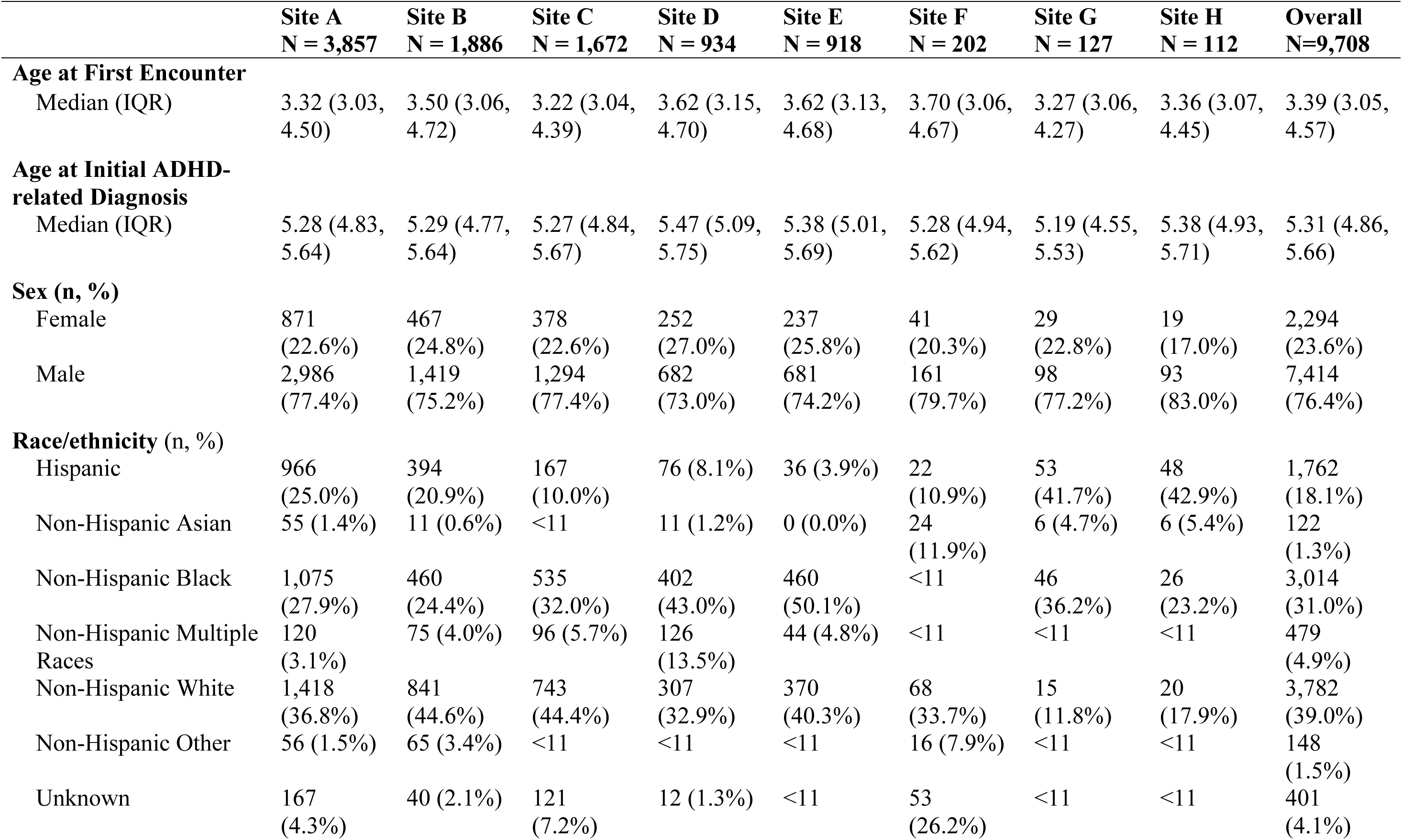
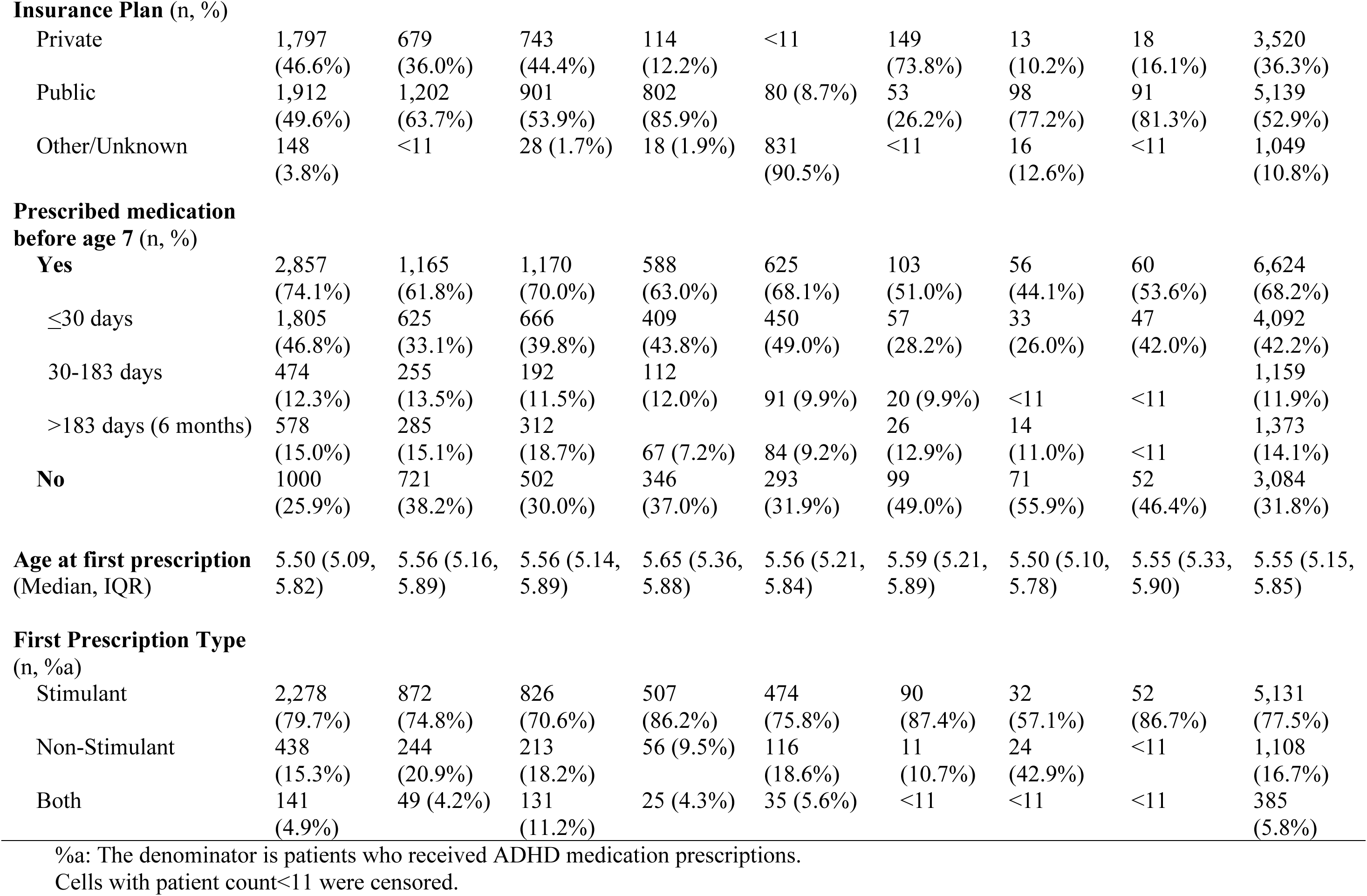
Patient demographics and clinical characteristics of children ages 3-5 years with ADHD (n=9,708).

### Timing of Medication Treatment

Of 9,708 children ages 3-5 years with ADHD, 42.2% (n=4,092) were prescribed medications within a month of the initial ADHD-related diagnosis (disorder-level or symptom-level) compared to 14.1% (n=1,373) prescribed >6 months after the initial diagnosis (**Table 1**). The median time from diagnosis to prescription varied by the child’s age at time of initial ADHD-related diagnosis: 391 days in 3-year-olds (n=264), 28 days in 4-year-olds (n=1,550), and 0 days in 5-year-olds (n=4,810) (**eTable 6**). The time from diagnosis to prescription varied widely across racial/ethnic subgroups (**Figure 2**). At or shortly after diagnosis (0-30 days), the cumulative incidence of medication prescription among ADHD patients was highest for Multiple Races, Non-Hispanic White, and Non-Hispanic Black patients (47.7%, 43.9%, and 41.8%, respectively), and lower for Hispanic (35.8%) and Non-Hispanic Asian patients (28.6%). At 2 years after diagnosis (450 person-years), prescription rates were high for Non-Hispanic White and Multiple Races patients (78.2% and 83.2%), lower for non-Hispanic Black (72.2%) and Hispanic patients (67.8%) and substantially lower for non-Hispanic Asian patients (55.6%).

**Figure 2.**
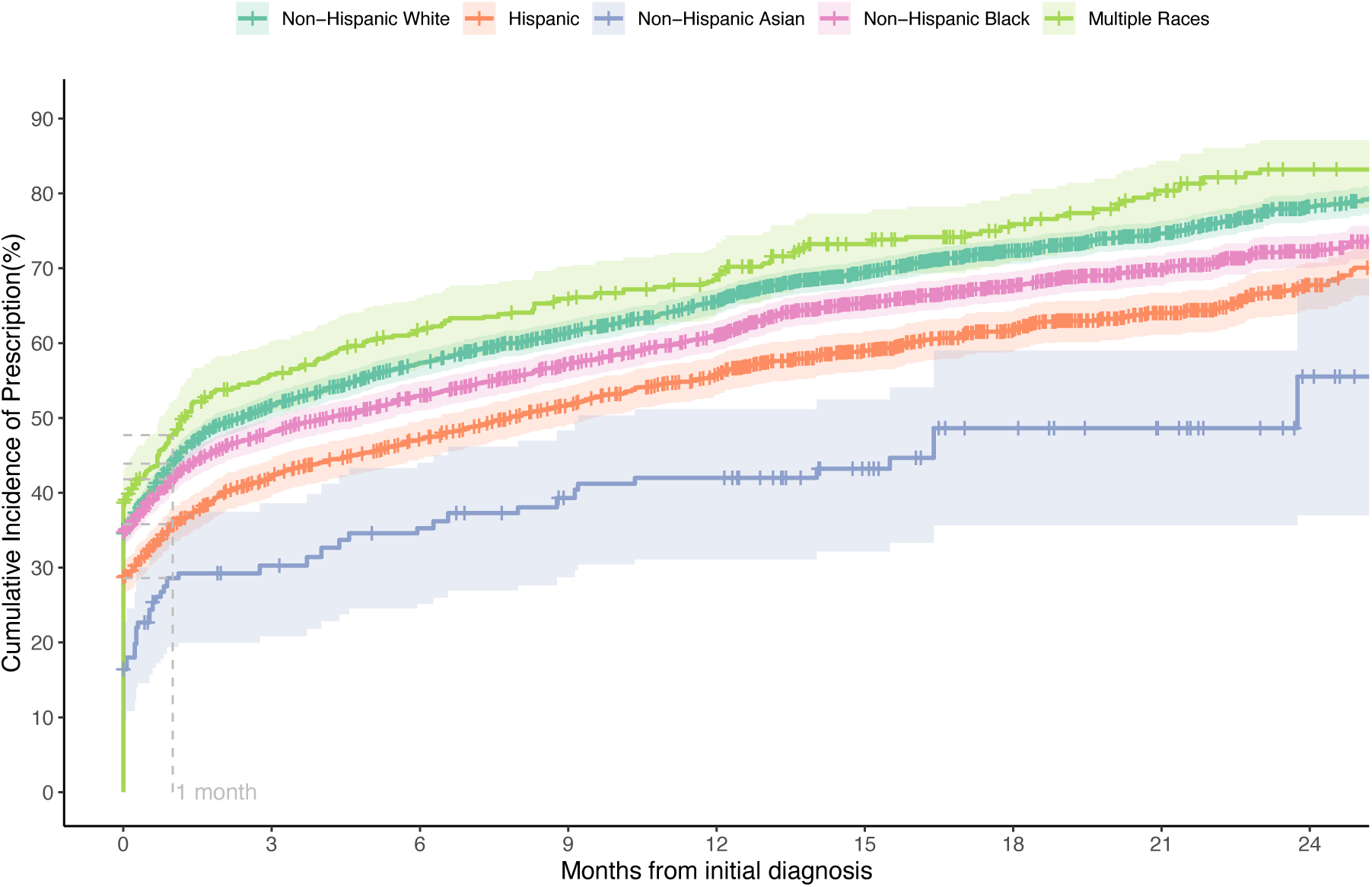
Time from initial diagnosis to prescription by race/ethnicity.* *Analysis excluded patients with other/unknown race or ethnicity. Maximum time to prescription was 47 months; x axis limited to 24 months for visual clarity.

### Regression Model

Cox regression model results examining time to first medication prescription after initial diagnosis and before age 7 are presented in **Figure 3**. After adjusting for year and type of diagnosis, comorbid conditions, primary care utilization, and institution, Asian (aHR 0.50, CI 0.38-0.65), Hispanic (aHR 0.75, CI 0.70-0.81), and Black (aHR 0.90, CI 0.85-0.96) patients with ADHD were less likely to be prescribed medication early compared to non-Hispanic White patients. Older (aHR 1.64, CI 1.57-1.72), male (aHR 1.17, CI 1.10-1.24) and publicly insured (aHR 1.10, CI 1.04-1.17) patients were more likely to be prescribed medication early compared to younger, female and privately insured patients, respectively.

**Figure 3.**
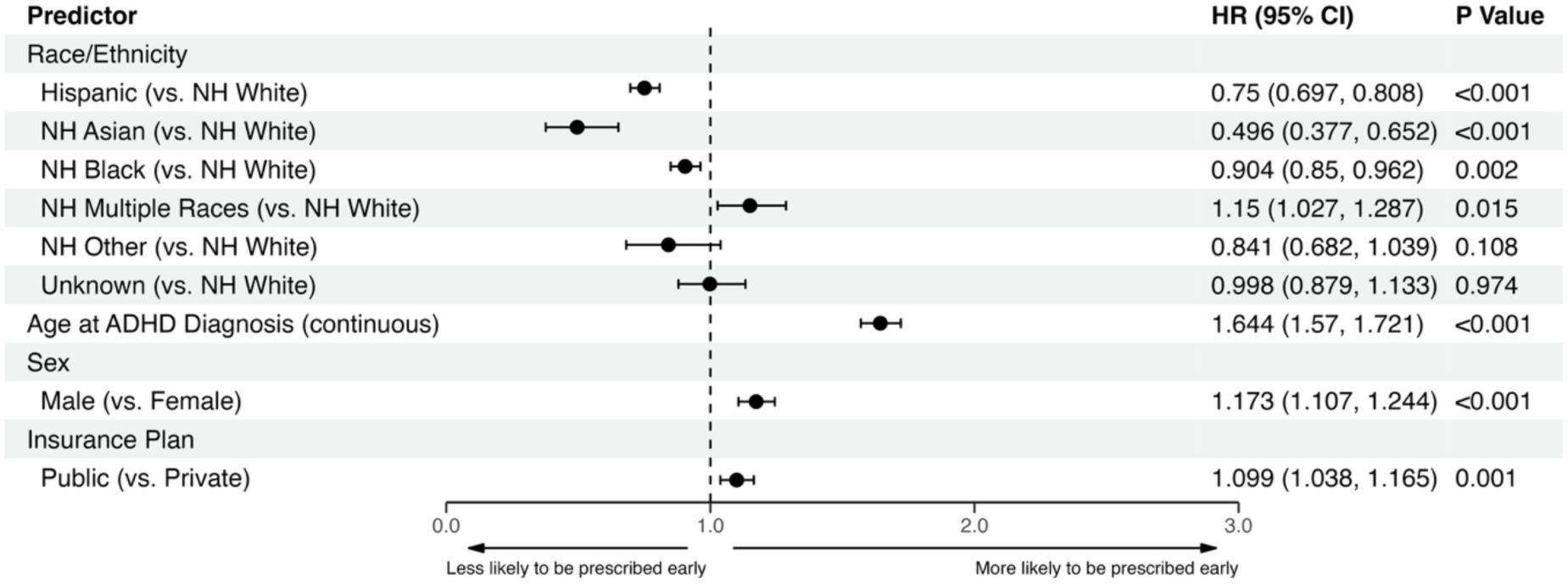
Time from initial diagnosis to prescription; adjusted cox regression model results.* * Adjusted for year of diagnosis, initial diagnosis type (disorder-level vs. symptom-level), primary care utilization (low: <5 visits, medium: 5-10 visits, high: >10 visits), presence of comorbidities, and institution. NH = Non-Hispanic.

### Secondary Outcomes

Of 6,624 preschool-age children prescribed ADHD medications, 40.5% (n= 2684) had an ADHD-related encounter within 2 months of the first prescription (25.9%-56.0% across institutions). Follow up visits were mostly in-person and less commonly via telehealth (**eFigure 1**). Data on rate of patient follow up via telephone or secure messaging was not available.

PCPs documented comorbid conditions, as recommended by practice guidelines, in 65.0% of patients with low variability (coefficient of variation=7.44%) across institutions (**Figure 4**). The most common comorbid conditions documented were language delay/disorder (33.3%), sleep problems (19.4%), and disruptive behavior disorders (19.2%).

**Figure 4.**
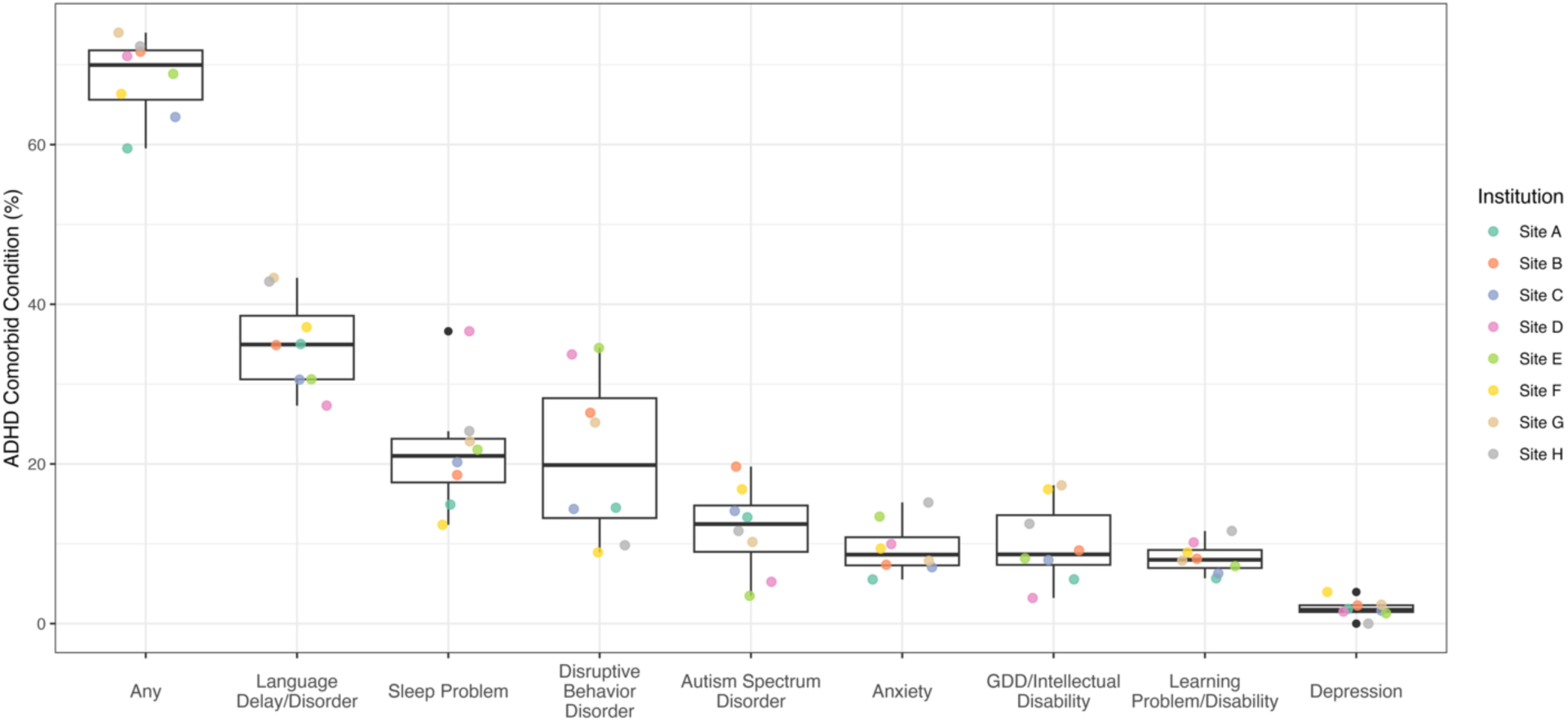
Documented developmental/behavioral comorbid conditions among children ages 3-5 years with ADHD. GDD = Global Developmental Delay

### Sensitivity analysis

When including non-specific diagnosis codes of behavior problems as the first ADHD-related concern, 27.8% of patients were prescribed within one month of first concern, while 25.9% were prescribed after > 6 months. The average time from initial concern to ADHD diagnosis was 3.4 months. A paired Wilcoxon test comparing time from behavior concern to prescription versus ADHD-related diagnosis to prescription showed a significant difference (p < 0.001). Among 7,730 children whose first ADHD-related diagnosis was disorder-level, 47.7% were prescribed within 30 days, compared to 22.9% of those whose initial diagnosis was symptom-level. The cumulative incidence curves crossed after one year (log-rank p=0.019), showing different prescribing trajectories based on diagnosis type (**eFigure 2**). Excluding one institution that had high rates of missing insurance data did not change the results substantively (data not shown).

## DISCUSSION

This study revealed high variability across large US pediatric health systems in rates of identification and medication treatment of children aged 3-5 years with ADHD. Overall, about two-thirds (68%) of preschool-age children with ADHD seen in primary care were prescribed ADHD medications before age 7, with over 40% of children prescribed within 30 days of their initial ADHD-related diagnosis – contrary to practice guidelines that recommend first starting with evidence-based non-pharmacological intervention. We found evidence for disparities in care with high rates of early medication treatment among White children and those with public insurance. These findings highlight the need to investigate factors influencing early medication treatment of preschoolers with ADHD, especially in specific subgroups of patients.

The prevalence of ADHD diagnosis varied across the 8 pediatric health systems in this study, aligning with CDC survey-based data that reported high variation in ADHD prevalence across US states.^24^ We found low rates of ADHD diagnosis in Hispanic, Asian, and Black children compared to White children, and high rates of diagnosis in publicly insured patients compared to privately insured patients, following previously reported trends.^12,13,20,25^

The high variation we found across institutions in the rate and timing of ADHD medication prescriptions highlights differences in clinical practice across health systems in the treatment of preschoolers with ADHD. Overall, medication prescription rates (ranging from 44.1% to 74.1% of patients) were higher than we had expected. While no clear benchmark exists for an appropriate rate of children 6 years and under with ADHD who should receive medications, clinical practice guidelines recommend medications as second-line treatment in cases with significant dysfunction or lack of response to behavioral treatment approaches. The rate of preschool-age children with ADHD prescribed medication by PCPs in the current study (68%) was higher than the rate prescribed medications by subspecialists in a prior study (46%), even though subspecialists often manage children with complex or severe presentations.^26^ Previous studies also reported that receipt of medication treatment was more common than first-line behavior treatment in US preschoolers.^17,20,27,28^ Future studies should investigate whether differences in clinical presentation, PCP knowledge of and patient access to non-pharmacological interventions, or family and PCP preferences are driving high rates of medication treatment.

Among children diagnosed with ADHD at ages 3-5 years, 42% were prescribed an ADHD medication within 30 days of the initial ADHD-related diagnosis (including symptom-level descriptors). Hence, over a third of patients lacked sufficient time for an evidence-based behavioral treatment prior to starting medications. This finding that, to our knowledge, has not been previously reported highlights the need to investigate factors that influence early medication prescriptions. In a sensitivity analysis that used non-specific behavior problems as the first indication of ADHD-related concern, rates of early prescription were lower (28%), suggesting that expanding the scope of investigation beyond diagnosis codes is necessary. Analyzing clinical free text documentation may enhance the accuracy of outlining treatment timelines of young children with ADHD, including timing of first concern, severity of symptoms, and recommendations for non-pharmacological interventions.

The high rates of early prescription that we found in both White children and in those with public insurance suggests that multiple factors influence the timing of medication prescriptions. Prior literature found that minoritized racial/ethnic groups are less likely to initiate ADHD medication treatment compared to White patients due to negative attitudes about ADHD medications; however, these same groups are also less likely to initiate and engage in parent behavioral training.^29,30^ Publicly insured patients often encounter systemic barriers to accessing subspecialists and evidence-based parent behavioral training,^30,31^ which may drive the clinician and the family to try medications early. Future interventions must address family-, clinician-, and system-level barriers to non-pharmacological treatments that may contribute, especially in certain sub-populations, to high rates of pharmacological treatment and low rates of non-pharmacological treatment of young children with ADHD.

We have also contributed to the limited literature on current rates of prescribing non-stimulants to treat preschool ADHD in primary care. In half of the examined health systems over 1 in 5 patients prescribed ADHD medications were prescribed non-stimulants (overall 21.1% of prescribed patients). Though there is limited evidence of the efficacy of non-stimulants in children under 6, prior studies found that subspecialists prescribed nonstimulants to about 1/3 of preschool-age children with ADHD, somewhat higher than PCP prescription rate in the present study.^26,32^

The proportion of preschoolers with ADHD who were seen for follow-up within 2 months of medication prescription varied across institutions from 25.9% to 56.0%. A prior single-site study found a similarly low rate of follow-up in preschoolers prescribed ADHD medications.^11^ However, these rates are likely an underestimate of actual follow-up rates. As previously reported, children with ADHD are frequently followed in primary care by telephone or secure messaging – a method that has been shown to contribute to achieving symptom improvement without an in-person or video-based visit.^33^ In addition, children may be referred to subspecialists who assume responsibility for follow-up care. Rates of documentation of comorbid conditions was high (approximately two-thirds of patients), suggesting PCPs adhere to guidelines in detecting common comorbid conditions in young children with ADHD.

### Limitations

Several limitations should be acknowledged. Study cohort identification was based on diagnostic codes, which carries the potential for misclassification error. To minimize this concern, we included in our cohort children who had at least one disorder-level ADHD diagnosis after age 4. We recognize that although we included symptom-level diagnoses of ADHD as the initial diagnosis, in some cases that first diagnosis may not represent the first time that concerns were raised regarding ADHD symptoms. In some patients, non-stimulant medications may have been prescribed to address sleep problems rather than ADHD symptoms. Our study focused on primary care management and did not include information on care provided by subspecialists (e.g., child psychiatrists, developmental-behavioral pediatricians). Finally, information on the rate of PCP recommendation for first-line behavioral treatment, typically documented as free text, was not available to us.^28,34^ Therefore, we could not determine if in some cases behavioral treatment was not available and, as recommended in guidelines, the PCP and family chose early medication treatment after weighing the risks and benefits. Future studies will need to extract information from clinical notes, which can be facilitated by natural language processing,^35,36^ to capture PCP recommendations about behavioral treatments, clinical reasoning behind early prescriptions, and the involvement of subspecialists in medication decisions.

## CONCLUSION

This large multi-site study revealed high variation in the identification and treatment of ADHD in children ages 3-5 years who presented in primary care settings. The high rate of medication prescriptions in preschool-age children with ADHD and the lack of delay between initial diagnosis and prescription require further investigation to assess the appropriateness of early medication treatment of young children. The variation in care among minoritized racial/ethnic groups uncovers disparities in care that require further exploration. Future studies will assess factors influencing prescribing patterns, including PCP and family preferences, severity of symptoms, and access to first-line non-pharmacological treatment across patient subgroups.

## Supporting information

eSupplement

## Data Availability

The datasets generated and analyzed in the current study contain protected patient health information and are therefore not publicly available.

## Acknowledgments

The research reported in this publication was conducted using PEDSnet, A Pediatric Clinical Research Network. PEDSnet has been developed with funding from the Patient-Centered Outcomes Research Institute (PCORI); PEDSnet’s participation in PCORnet is funded through PCORI award RI-CHOP-01-PS1. This publication includes data from the following PEDSnet institutions: Children’s Hospital of Colorado, Children’s Hospital of Philadelphia, Cincinnati Children’s Hospital Medical Center, Lurie Children’s Hospital of Chicago, Nationwide Children’s Hospital, Nemours Children’s Health, Packard Children’s Hospital Stanford, Texas Children’s Hospital.

## Conflict of Interest Disclosures (includes financial disclosures)

Dr. Wallis has been paid as a consultant to *Healthy Steps* and *Thomas Jefferson University*. She serves as an expert witness for Kline Spector, LLC. All other authors have no conflicts of interest to disclose.

## Contributorship Statement

Dr. Bannett conceptualized and designed the study, defined and coordinated data extraction, participated in data analyses and drafting the manuscript, and reviewed and revised the manuscript.

Ms. Luo participated in study design, carried out the data analyses, participated in drafting the manuscript, and reviewed and revised the manuscript.

Mr. Azuero-dajud participated in study design, data extraction and analyses, and critically reviewed and revised the manuscript.

Drs. Feldman and Forrest participated in conceptualization of the study, interpretation of the data, and critically reviewed and revised the manuscript.

Drs. Brink, Froehlich, Harris, Kan, Wallis, Whelan, and Spector participated in interpretation of the data, and critically reviewed and revised the manuscript.

Ms. Luo and Mr. Azuero-dajud had full access to all the data in the study and take responsibility for the integrity of the data and the accuracy of the data analysis.

All authors approved the final manuscript as submitted and are responsible for all aspects of the work.

## References

1. Li Y, Yan X, Li Q, et al. Prevalence and Trends in Diagnosed ADHD Among US Children and Adolescents, 2017-2022. JAMA Netw Open. Oct 2 2023;6(10):e2336872. doi:10.1001/jamanetworkopen.2023.36872

2. Danielson ML, Claussen AH, Bitsko RH, et al. ADHD Prevalence Among U.S. Children and Adolescents in 2022: Diagnosis, Severity, Co-Occurring Disorders, and Treatment. J Clin Child Adolesc Psychol. May-Jun 2024;53(3):343–360. doi:10.1080/15374416.2024.2335625

3. Perrin HT, Heller NA, Loe IM. School Readiness in Preschoolers With Symptoms of Attention-Deficit/Hyperactivity Disorder. Pediatrics. Aug 2019;144(2):DOI: 10.1542/peds.2019-0038. doi:10.1542/peds.2019-0038

4. Spira EG, Fischel JE. The impact of preschool inattention, hyperactivity, and impulsivity on social and academic development: a review. Journal of child psychology and psychiatry, and allied disciplines. Jul 2005;46(7):755–73. doi:10.1111/j.1469-7610.2005.01466.x

5. Wolraich M, Brown L, Brown RT, et al. ADHD: clinical practice guideline for the diagnosis, evaluation, and treatment of attention-deficit/hyperactivity disorder in children and adolescents. Pediatrics. Nov 2011;128(5):1007–22. doi:10.1542/peds.2011-2654

6. Wolraich ML, Hagan JF, Jr., Allan C, et al. Clinical Practice Guideline for the Diagnosis, Evaluation, and Treatment of Attention-Deficit/Hyperactivity Disorder in Children and Adolescents. Pediatrics. Oct 2019;144(4)doi:10.1542/peds.2019-2528

7. Charach A, Carson P, Fox S, et al. Interventions for preschool children at high risk for ADHD: a comparative effectiveness review. Pediatrics. May 2013;131(5):e1584–604. doi:10.1542/peds.2012-0974

8. Visser SN, Bitsko RH, Danielson ML, et al. Treatment of Attention Deficit/Hyperactivity Disorder among Children with Special Health Care Needs. J Pediatr. Jun 2015;166(6):1423–30.e1-2. doi:10.1016/j.jpeds.2015.02.018

9. Albert M, Rui P, Ashman JJ. Physician Office Visits for Attention-deficit/Hyperactivity Disorder in Children and Adolescents Aged 4-17 Years: United States, 2012-2013. NCHS data brief. Jan 2017;(269):1–8.

10. Davis DW, Feygin Y, Creel L, et al. Epidemiology of Treatment for Preschoolers on Kentucky Medicaid Diagnosed with Attention-Deficit/Hyperactivity Disorder. Journal of child and adolescent psychopharmacology. Sep 2020;30(7):448–455. doi:10.1089/cap.2020.0015

11. Bannett Y, Feldman HM, Gardner RM, et al. Attention-Deficit/Hyperactivity Disorder in 2-to 5-Year-Olds: A Primary Care Network Experience. Academic pediatrics. Apr 28 2020;doi:10.1016/j.acap.2020.04.009

12. Danielson ML, Visser SN, Gleason MM, et al. A National Profile of Attention-Deficit Hyperactivity Disorder Diagnosis and Treatment Among US Children Aged 2 to 5 Years. J Dev Behav Pediatr. Sep 2017;38(7):455–464. doi:10.1097/dbp.0000000000000477

13. Girand HL, Litkowiec S, Sohn M. Attention-Deficit/Hyperactivity Disorder and Psychotropic Polypharmacy Prescribing Trends. Pediatrics. Jun 2 2020;doi:10.1542/peds.2019-2832

14. Fiks AG, Ross ME, Mayne SL, et al. Preschool ADHD Diagnosis and Stimulant Use Before and After the 2011 AAP Practice Guideline. Pediatrics. Dec 2016;138(6)doi:10.1542/peds.2016-2025

15. Danielson ML, Bohm MK, Newsome K, et al. Trends in Stimulant Prescription Fills Among Commercially Insured Children and Adults - United States, 2016-2021. MMWR Morbidity and mortality weekly report. Mar 31 2023;72(13):327–332. doi:10.15585/mmwr.mm7213a1

16. Hales CM, Kit BK, Gu Q, et al. Trends in Prescription Medication Use Among Children and Adolescents-United States, 1999-2014. Jama. May 15 2018;319(19):2009–2020. doi:10.1001/jama.2018.5690

17. Visser SN, Danielson ML, Wolraich ML, et al. Vital Signs: National and State-Specific Patterns of Attention Deficit/Hyperactivity Disorder Treatment Among Insured Children Aged 2-5 Years - United States, 2008-2014. MMWR Morbidity and mortality weekly report. May 6 2016;65(17):443–50. doi:10.15585/mmwr.mm6517e1

18. Kamimura-Nishimura KI, Epstein JN, Froehlich TE, et al. Factors Associated with Attention Deficit Hyperactivity Disorder Medication Use in Community Care Settings. J Pediatr. Oct 2019;213:155–162.e1. doi:10.1016/j.jpeds.2019.06.025

19. Walls M, Allen CG, Cabral H, et al. Receipt of Medication and Behavioral Therapy Among a National Sample of School-Age Children Diagnosed With Attention-Deficit/Hyperactivity Disorder. Academic pediatrics. Apr 2018;18(3):256–265. doi:10.1016/j.acap.2017.10.003

20. Shi Y, Hunter Guevara LR, Dykhoff HJ, et al. Racial Disparities in Diagnosis of Attention-Deficit/Hyperactivity Disorder in a US National Birth Cohort. JAMA Network Open. 2021;4(3):e210321–e210321. doi:10.1001/jamanetworkopen.2021.0321

21. Lindly OJ, Nasol E, Tarazi CL, et al. Toward Equitable Health Outcomes for Diverse Children With ADHD and Their Families. Academic pediatrics. Feb 3 2021;doi:10.1016/j.acap.2021.01.020

22. Coker TR, Elliott MN, Toomey SL, et al. Racial and Ethnic Disparities in ADHD Diagnosis and Treatment. Pediatrics. Sep 2016;138(3)doi:10.1542/peds.2016-0407

23. Vandenbroucke JP, von Elm E, Altman DG, et al. Strengthening the Reporting of Observational Studies in Epidemiology (STROBE): explanation and elaboration. PLoS Med. Oct 16 2007;4(10):e297. doi:10.1371/journal.pmed.0040297

24. Danielson ML, Holbrook JR, Bitsko RH, et al. State-Level Estimates of the Prevalence of Parent-Reported ADHD Diagnosis and Treatment Among U.S. Children and Adolescents, 2016 to 2019. Journal of attention disorders. Nov 2022;26(13):1685–1697. doi:10.1177/10870547221099961

25. Bax AC, Bard DE, Cuffe SP, et al. The Association Between Race/Ethnicity and Socioeconomic Factors and the Diagnosis and Treatment of Children with Attention-Deficit Hyperactivity Disorder. J Dev Behav Pediatr. Feb/Mar 2019;40(2):81–91. doi:10.1097/dbp.0000000000000626

26. Blum NJ, Shults J, Harstad E, et al. Common Use of Stimulants and Alpha-2 Agonists to Treat Preschool Attention-Deficit Hyperactivity Disorder: A DBPNet Study. J Dev Behav Pediatr. Sep 2018;39(7):531–537. doi:10.1097/dbp.0000000000000585

27. Danielson ML, Visser SN, Chronis-Tuscano A, et al. A National Description of Treatment among United States Children and Adolescents with Attention-Deficit/Hyperactivity Disorder. J Pediatr. Jan 2018;192:240–246.e1. doi:10.1016/j.jpeds.2017.08.040

28. Bannett Y, Gardner RM, Posada J, et al. Rate of Pediatrician Recommendations for Behavioral Treatment for Preschoolers With Attention-Deficit/Hyperactivity Disorder Diagnosis or Related Symptoms. JAMA pediatrics. 2021;doi:10.1001/jamapediatrics.2021.4093

29. Kamimura-Nishimura K, Bush H, Amaya de Lopez P, et al. Understanding Barriers and Facilitators of Attention-Deficit/Hyperactivity Disorder Treatment Initiation and Adherence in Black and Latinx Children. Academic pediatrics. Aug 2023;23(6):1175–1186. doi:10.1016/j.acap.2023.03.014

30. Green CD, Langberg JM. A Review of Predictors of Psychosocial Service Utilization in Youth with Attention-Deficit/Hyperactivity Disorder. Clin Child Fam Psychol Rev. Jun 2022;25(2):356–375. doi:10.1007/s10567-021-00368-y

31. Bishop TF, Press MJ, Keyhani S, et al. Acceptance of insurance by psychiatrists and the implications for access to mental health care. JAMA Psychiatry. Feb 2014;71(2):176–81. doi:10.1001/jamapsychiatry.2013.2862

32. Harstad E, Shults J, Barbaresi W, et al. α2-Adrenergic Agonists or Stimulants for Preschool-Age Children With Attention-Deficit/Hyperactivity Disorder. Jama. May 25 2021;325(20):2067–2075. doi:10.1001/jama.2021.6118

33. Epstein JN, Kelleher KJ, Baum R, et al. Specific Components of Pediatricians’ Medication-Related Care Predict Attention-Deficit/Hyperactivity Disorder Symptom Improvement. Journal of the American Academy of Child and Adolescent Psychiatry. Jun 2017;56(6):483–490.e1. doi:10.1016/j.jaac.2017.03.014

34. Pillai M, Posada J, Gardner RM, et al. Measuring quality-of-care in treatment of young children with attention-deficit/hyperactivity disorder using pre-trained language models. Journal of the American Medical Informatics Association : JAMIA. Apr 3 2024;31(4):949–957. doi:10.1093/jamia/ocae001

35. Bannett Y, Bassett HK, Morse KE. Natural Language Processing: Set to Transform Pediatric Research. Hosp Pediatr. Jan 1 2025;15(1):e12–e14. doi:10.1542/hpeds.2024-008115

36. Bannett Y, Gunturkun F, Pillai M, et al. Applying Large Language Models to Assess Quality of Care: Monitoring ADHD Medication Side Effects. Pediatrics. Jan 1 2025;155(1)doi:10.1542/peds.2024-067223

