## Supplementary material for "Timing of Medication Treatment in Children 3-5-Years-old with ADHD: A PEDSnet Study": eSupplement

**e-Supplement.**

**eTable 1.** List of codes used for well care visits and immunizations.
**eTable 2.** List of codes used for ADHD diagnosis and ADHD medications.
**eTable 3.** List of codes used for developmental/behavioral comorbid conditions.
**eTable 4.** List of codes for behavioral problems used in sensitivity analysis.
**eTable 5.** Patient demographics of children seen in primary care at ages 3-5 years across 8 institutions (n=712,478)
**eTable 6.** Time from diagnosis to prescription among preschool-age children with ADHD, stratified by patient age at time of initial ADHD-related diagnosis.
**eFigure 1**. Follow up of children 3-5 years who were prescribed ADHD medications.
**eFigure 2.** Time from initial ADHD diagnosis to medication prescription, stratified by the type of initial ADHD diagnosis (symptom-level first vs. disorder-level first).

**eTable 1.** List of codes used for well care visits and immunizations.

| Concept id*/ CPT code | Name | Domain | Vocabulary |
|---|---|---|---|
| 4088016 | Child examination | Procedure | SNOMED |
| 1012596 | Immunization administration (includes percutaneous, intradermal, subcutaneous, or intramuscular injections) | Procedure | CPT4 |
| 1012599 | Immunization administration by intranasal or oral route | Procedure | CPT4 |
| 90620/1 | Meningococcal recombinant, serogroup B | Drug | CPT4 |
| 90633 | Hepatitis A vaccine, pediatric/adolescent | Drug | CPT4 |
| 90647/8 | Hemophilus influenza B vaccine (Hib) | Drug | CPT4 |
| 90651 | Human Papillomavirus vaccine (HPV) | Drug | CPT4 |
| 90670 | Pneumococcal conjugate vaccine, 13 valent | Drug | CPT4 |
| 90672 | Influenza virus vaccine, quadrivalent, live | Drug | CPT4 |
| 90680 | Rotavirus vaccine, pentavalent (Rotateq) | Drug | CPT4 |
| 90681 | Rotavirus vaccine, attenuated (Rotarix) | Drug | CPT4 |
| 90686 | Influenza virus vaccine, quadrivalent | Drug | CPT4 |
| 90696 | Diphtheria, tetanus toxoids, acellular pertussis vaccine and poliovirus vaccine, inactivated (DTaP-IPV) | Drug | CPT4 |
| 90698 | Diphtheria, tetanus toxoids, acellular pertussis vaccine, hemophilus influenza Type B, and poliovirus vaccine, inactivated (DTaP-HIB-IPV) | Drug | CPT4 |
| 90700 | diphtheria, tetanus toxoids, acellular pertussis vaccine (DtaP) | Drug | CPT4 |
| 90707 | Measles, mumps, and rubella vaccine (MMR) | Drug | CPT4 |
| 90710 | Measles, mumps, rubella, and varicella vaccine (MMRV) | Drug | CPT4 |
| 90713 | Poliovirus vaccine, inactivated (IPV) | Drug | CPT4 |
| 90714 | Tetanus and diphtheria toxoids (Td) | Drug | CPT4 |
| 90715 | Tetanus, diphtheria toxoids and acellular pertussis vaccine (Tdap) | Drug | CPT4 |
| 90716 | Varicella virus vaccine, live | Drug | CPT4 |
| 90723 | Diphtheria, tetanus toxoids, acellular pertussis vaccine, Hepatitis B, and poliovirus vaccine, inactivated (DtaP-HepB-IPV) | Drug | CPT4 |
| 90732 | Pneumococcal polysaccharide vaccine, 23-valent | Drug | CPT4 |
| 90734 | Meningicoccal conjugate vaccine, serogoups A, C, Y and W-135 (quadtivalent) | Drug | CPT4 |
| 90744 | Hepatitis B vaccine, pediatric/adolescent | Drug | CPT4 |

*We included these concept ids and all of their descendants
CPT= Current Procedural Terminology

**eTable 2.** List of codes used for ADHD diagnosis and ADHD medications.

| Condition type | ICD-10 code | Concept id* | Condition/Medication name |
|---|---|---|---|
| **ADHD diagnosis** | | | |
| ADHD (disorder-level) | F90.0 | 35207262 | Attention-deficit hyperactivity disorder, predominantly inattentive type |
| ADHD (disorder-level) | F90.1 | 35207263 | Attention-deficit hyperactivity disorder, predominantly hyperactive type |
| ADHD (disorder-level) | F90.2 | 45552506 | Attention-deficit hyperactivity disorder, combined type |
| ADHD (disorder-level) | F90.8 | 35207264 | Attention-deficit hyperactivity disorder, other type |
| ADHD (disorder-level) | F90.9 | 35207265 | Attention-deficit hyperactivity disorder, unspecified type |
| ADHD (symptom-level) | F90.9 | 45533124 | Hyperactivity; Hyperkinesis |
| ADHD (symptom-level) | R41.840 | 45582710 | Inattention; Attention and concentration deficit |
| ADHD (symptom-level) | R41.844 | 45539344 | Executive function deficit |
| ADHD (symptom-level) | R45.87 | 45568136 | Impulsiveness |
| **ADHD medications** | | | |
| **Stimulants** | | | |
| | | 705944 | Methylphenidate |
| | | 731533 | Dexmethylphenidate |
| | | 714785 | Amphetamine |
| | | 719311 | Dextroamphetamine |
| | | 709567 | Lisdexamfetamine |
| **Non-stimulants** | | | |
| | | 1344965 | Guanfacine |
| | | 1398937 | Clonidine |
| | | 742185 | Atomoxetine |

*We included these concept ids and all of their descendants

**eTable 3.** List of codes used for developmental/behavioral comorbid conditions.

| Condition type | ICD-10 code | Condition name |
|---|---|---|
| Autism | F84.0 | Autism spectrum disorder |
| Autism | F84.5 | Asperger's syndrome |
| Autism | F84.8 | Other pervasive developmental disorders |
| Autism | F84.9 | Pervasive developmental disorder, unspecified |
| Autism | F94.8 | Other childhood disorders of social functioning |
| Autism | F94.9 | Childhood disorder of social functioning, unspecified |
| Autism | F80.82 | Social pragmatic communication disorder |
| Anxiety | F40 | Phobic anxiety disorders |
| Anxiety | F41 | Other anxiety disorders |
| Anxiety | F93.0 | Separation anxiety disorder of childhood |
| Anxiety | F94.0 | Selective mutism |
| Depression | F32 | Depressive episode |
| Depression | F33 | Recurrent depressive disorder |
| Depression | F34 | Persistent mood [affective] disorder |
| Depression | F39 | Unspecified mood [affective] disorder |
| Disruptive behavior disorder | F91 | Conduct disorder, Oppositional defiant disorder |
| Disruptive behavior disorder | F63.81 | Intermittent Explosive Disorder |
| Global Developmental Delay/Intellectual disability | F88 | Global Developmental Delay/Other disorders of psychological development |
| Global Developmental Delay/Intellectual disability | F89 | Unspecified disorder of psychological development |
| Global Developmental Delay/Intellectual disability | F70 | Mild intellectual disabilities |
| Global Developmental Delay/Intellectual disability | F71 | Moderate intellectual disabilities |
| Global Developmental Delay/Intellectual disability | F72 | Severe intellectual disabilities |
| Global Developmental Delay/Intellectual disability | F73 | Profound intellectual disabilities |
| Global Developmental Delay/Intellectual disability | F79 | Unspecified intellectual disabilities |
| Learning problem/ disability | F81 | Specific developmental disorders of scholastic skills |
| Learning problem/ disability | Z55 | Problems related to education and literacy |
| Learning problem/ disability | R48.0 | Dyslexia and alexia |

| | | |
|---|---|---|
| Language delay/ disorder | F80.0 | Phonological disorder |
| Language delay/ disorder | F80.1 | Expressive language disorder |
| Language delay/ disorder | F80.2 | Mixed receptive-expressive language disorder |
| Language delay/ disorder | F80.81 | Childhood onset fluency disorder |
| Language delay/ disorder | F80.89 | Other developmental disorders of speech and language |
| Language delay/ disorder | F80.9 | Developmental disorder of speech and language, unspecified |
| Language delay/ disorder | R47.89 | Other speech disturbances |
| Language delay/ disorder | R47.9 | Unspecified speech disturbances |
| Sleep problems | G47 | Sleep disorders |
| Sleep problems | F51 | Sleep disorders not due to a substance or known physiological condition |
| Sleep problems | Z72.82 | Problems related to sleep |
| Sleep problems | Z73.81 | Behavioral insomnia of childhood |

**eTable 4.** List of codes for behavioral problems used in sensitivity analysis.

| Condition type | ICD-10 code | Condition name |
|---|---|---|
| Behavioral problem | F91 | Temper tantrums / oppositional behavior / disruptive behavior |
| Behavioral problem | R45.4 | Irritability / anger |
| Behavioral problem | R45.5 | Aggressive outburst / Hostility |
| Behavioral problem | R45.6 | Violent behavior |
| Behavioral problem | R46.89 | Behavior concern / behavior problem in child / aggression |
| Behavioral problem | Z72.810 | Child and adolescent antisocial behavior, behavior problem at school |

**eTable 5.** Patient demographics of children seen in primary care at ages 3-5 years across 8 institutions (n=712,478)

| | Site A<br>N = 310,150 | Site B<br>N = 95,619 | Site C<br>N = 159,989 | Site D<br>N = 56,714 | Site E<br>N = 29,837 | Site F<br>N = 44,152 | Site G<br>N = 5,041 | Site H<br>N = 10,976 | Overall<br>N=712,478 |
|---|---|---|---|---|---|---|---|---|---|
| **Age at First Encounter** | | | | | | | | | |
| Median (IQR) | 3.18 (3.03, 4.09) | 3.25 (3.04, 4.23) | 3.17 (3.04, 4.02) | 3.47 (3.12, 4.28) | 3.44 (3.11, 4.34) | 3.27 (3.04, 4.38) | 3.39 (3.08, 4.45) | 3.29 (3.08, 4.07) | 3.22 (3.04, 4.13) |
| **Sex (n, %)** | | | | | | | | | |
| Female | 151,271 (48.8%) | 46,234 (48.4%) | 77,317 (48.3%) | 27,488 (48.5%) | 14,568 (48.8%) | 21,444 (48.6%) | 2,344 (46.5%) | 5,213 (47.5%) | 345,879 (48.5%) |
| Male | 158,879 (51.2%) | 49,385 (51.6%) | 82,672 (51.7%) | 29,226 (51.5%) | 15,269 (51.2%) | 22,708 (51.4%) | 2,697 (53.5%) | 5,763 (52.5%) | 366,599 (51.5%) |
| **Race/ethnicity (n, %)** | | | | | | | | | |
| Hispanic | 104,703 (33.8%) | 18,342 (19.2%) | 15,338 (9.6%) | 8,678 (15.3%) | 1,855 (6.2%) | 4,871 (11.0%) | 2,047 (40.6%) | 4,999 (45.5%) | 160,833 (22.6%) |
| Non-Hispanic Asian | 20,446 (6.6%) | 4,601 (4.8%) | 8,627 (5.4%) | 4,135 (7.3%) | 658 (2.2%) | 8,424 (19.1%) | 318 (6.3%) | 531 (4.8%) | 47,740 (6.7%) |
| Non-Hispanic Black | 46,732 (15.1%) | 21,875 (22.9%) | 40,986 (25.6%) | 30,196 (53.2%) | 15,940 (53.4%) | 1,179 (2.7%) | 1,651 (32.8%) | 3,043 (27.7%) | 161,602 (22.7%) |
| Non-Hispanic Multiple Races | 8,395 (2.7%) | 3,214 (3.4%) | 6,177 (3.9%) | 4,292 (7.6%) | 1,416 (4.7%) | 1,562 (3.5%) | 139 (2.8%) | 511 (4.7%) | 25,706 (3.6%) |
| Non-Hispanic White | 102,720 (33.1%) | 40,308 (42.2%) | 73,588 (46.0%) | 8,701 (15.3%) | 9,375 (31.4%) | 10,109 (22.9%) | 591 (11.7%) | 1,284 (11.7%) | 246,676 (34.6%) |
| Non-Hispanic Other | 6,940 (2.2%) | 4,343 (4.5%) | 195 (0.1%) | 91 (0.2%) | 310 (1.0%) | 3,909 (8.9%) | 229 (4.5%) | 475 (4.3%) | 16,492 (2.3%) |
| Unknown | 20,214 (6.5%) | 2,936 (3.1%) | 15,078 (9.4%) | 621 (1.1%) | 283 (0.9%) | 14,098 (31.9%) | 66 (1.3%) | 133 (1.2%) | 53,429 (7.5%) |
| **Insurance Plan (n, %)** | | | | | | | | | |
| Private | 181,751 (58.6%) | 53,362 (55.8%) | 97,798 (61.1%) | 8,257 (14.6%) | 234 (0.8%) | 36,862 (83.5%) | 859 (17.0%) | 1,345 (12.3%) | 380,468 (53.4%) |
| | 106,068 (34.2%) | 41,486 (43.4%) | 57,127 (35.7%) | 47,081 (83.0%) | 1,826 (6.1%) | 7,290 (16.5%) | 3,445 (68.3%) | 9,222 (84.0%) | 273,545 (38.4%) |

| | | | | | | | | | |
|---|---|---|---|---|---|---|---|---|---|
| Public | 22,331 (7.2%) | 771 (0.8%) | 5,064 (3.2%) | 1,376 (2.4%) | 27,777 (93.1%) | 0 (0.0%) | 737 (14.6%) | 409 (3.7%) | 58,465 (8.2%) |
| Other/Unknown | 181,751 (58.6%) | 53,362 (55.8%) | 97,798 (61.1%) | 8,257 (14.6%) | 234 (0.8%) | 36,862 (83.5%) | 859 (17.0%) | 1,345 (12.3%) | 380,468 (53.4%) |
| **Patients with at least 1 ADHD diagnosis at age 4-5 years (n, %)** | 3,857 (1.2%) | 1,886 (2.0%) | 1,672 (1.0%) | 934 (1.6%) | 918 (3.1%) | 202 (0.5%) | 127 (2.5%) | 112 (1.0%) | 9,708 (1.4%) |

**eTable 6.** Time from diagnosis to prescription among preschool-age children with ADHD, stratified by patient age at time of initial ADHD-related diagnosis.

| | Age at initial ADHD diagnosis | | | |
|---|---|---|---|---|
| | **3 Years** (N=376) | **4 Years** (N=2405) | **5 Years** (N=6927) | **Overall** (N=9708) |
| **Prescribed medication before age 7** (n, %) | | | | |
| **Yes** | | | | |
| Median (IQR) in days | 390.5 (204.2, 672.0) | 28.0 (0.0, 289.2) | 0.0 (0.0, 55.0) | 2.0 (0.0, 127.0) |
| ≤30 days | 6 (1.6%) | 785 (32.6%) | 3301 (47.7%) | 4092 (42.2%) |
| 30-183 days | 55 (14.6%) | 238 (9.9%) | 866 (12.5%) | 1159 (11.9%) |
| >183 days (6 months) | 203 (54.0%) | 527 (21.9%) | 643 (9.3%) | 1373 (14.1%) |
| **No** | 112 (29.8%) | 855 (35.6%) | 2117 (30.6%) | 3084 (31.8%) |

**eFigure 1**. Follow up of children 3-5 years who were prescribed ADHD medications (n=6624).

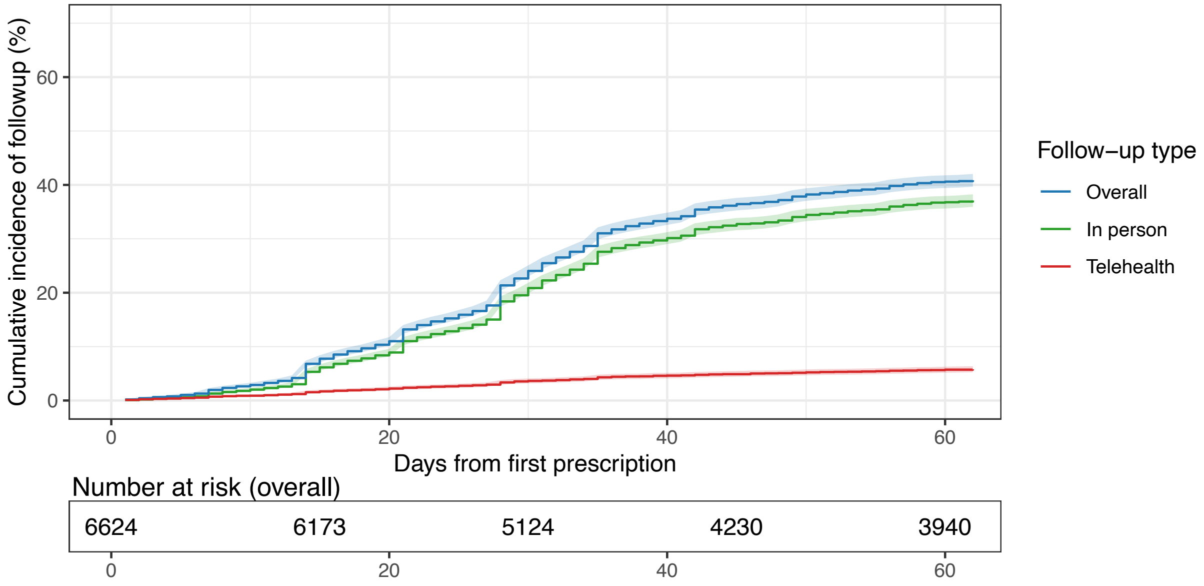

**eFigure 2.** Time from initial ADHD diagnosis to medication prescription, stratified by the type of initial ADHD diagnosis (symptom-level first vs. disorder-level first).

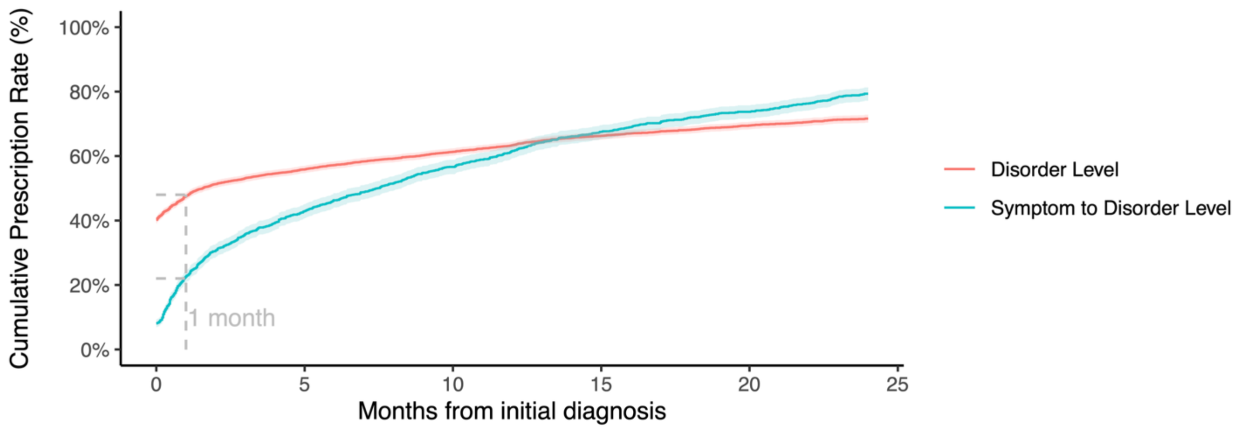
